# Who Is Reached by Supervised Psilocybin Services? Oregon Services Versus National Psilocybin Use

**DOI:** 10.64898/2026.09.03.26362170

**Authors:** Gabriel P. A. Costa, Christina Riggione, Christopher Pittenger, Joao P. De Aquino

**Affiliations:** Department of Psychiatry, Yale University School of Medicine, New Haven, CT, United States; Clinical Neuroscience Research Unit (CNRU), Connecticut Mental Health Center (CMHC), New Haven, CT, United States; VA Connecticut Healthcare System, West Haven, CT, USA

**Author notes:** **Corresponding Authors:** Joao P. De Aquino, M.D. Department of Psychiatry Yale School of Medicine, 34 Park St, 333B, New Haven, CT 06519, Gabriel P. A. Costa, M.D. Department of Psychiatry Yale School of Medicine, 34 Park St, 332, New Haven, CT 06519.

**Keywords:** psilocybin, psychedelics, Oregon Psilocybin Services, NSDUH, substance use disorders, policy

## Abstract

Supervised psilocybin services are expanding across the United States, yet how individuals accessing these programs compare with those using psilocybin in the general population is unknown. We conducted a cross-sectional descriptive comparison of Oregon Psilocybin Services (OPS) data from licensed service centers in 2025 (5,935 encounters) and the 2024 National Survey on Drug Use and Health (NSDUH) public-use file (1,822 of 47,299 adults reporting past-year psilocybin use), examining demographics, reasons for use, self-reported disability, and past-year psychiatric conditions and treatment. OPS clients were predominantly female (58.2%) and aged 35 or older (82.1%), whereas adults reporting psilocybin use nationally were predominantly male (64.0%) and younger than 35 (56.7%). Racial and ethnic minority representation was narrower in Oregon (0.9% Black, 2.5% Hispanic vs. 5.4% and 14.3% nationally). OPS clients reported higher incomes (58.5% earning >$95,000) and primarily cited wellness; self-reported disability was uncommon. Adults using psilocybin nationally had high past-year rates of major depressive episode (22.7%), serious psychological distress (36.8%), substance use disorder (61.7%), and mental health treatment receipt (42.9%). Oregon’s open-access model selects higher-income, clinically stable, wellness-oriented adults, while millions with psychiatric and substance-use burden use psilocybin outside supervised channels; its safety and utilization data should not be generalized.

## INTRODUCTION

The U.S. is rapidly changing how it regulates psychedelics. Psilocybin has been at the center of this shift: more than 30 states have considered new legislation to expand legal access. (Siegel et al., 2023; University of California. Berkeley Center for the Science of Psychedelics, 2026) In 2023, Oregon became the first state to implement supervised psilocybin services. (“Oregon Revised Statutes Chapter 475A: Psilocybin Services (Ballot Measure 109),” 2020) Oregon’s model requires no clinical diagnosis; services are open to adults aged 21 years or older, subject to brief safety screening that excludes current lithium use, active thoughts of harm to self or others, and a history of active psychosis. (“Oregon Revised Statutes Chapter 475A: Psilocybin Services (Ballot Measure 109),” 2020; Oregon Secretary of State, 2026) Clients complete a required preparation session, a supervised administration session at a licensed service center, and an optional integration session. Facilitators use a nondirective approach and may not provide psychotherapy or medical care. (“Oregon Revised Statutes Chapter 475A: Psilocybin Services (Ballot Measure 109),” 2020; Oregon Secretary of State, 2026) Psilocybin products are manufactured, tested, and dispensed within the licensed system, and clients cannot supply their own. These services are the only legal route to psilocybin in Oregon, with possession outside the program remaining prohibited under state law. (“Oregon Revised Statutes Chapter 475A: Psilocybin Services (Ballot Measure 109),” 2020) Colorado has since launched a similar open-access program, and several other states have introduced legislation that would allow psilocybin access without requiring a clinical diagnosis. (University of California. Berkeley Center for the Science of Psychedelics, 2026) Meanwhile, unsupervised psilocybin use has increased nationwide. Almost 8 million individuals aged 12 or older reported past-year use in 2024, (Yang et al., 2026) and state-level decriminalization has been associated with further increase. (Black et al., 2026) Initial descriptive reports of Oregon’s service utilization have appeared, (Cheung et al., 2026; Yu et al., 2026) yet who supervised services reach relative to the broader population using psilocybin nationally has not been examined.

Two recently available data sources allow this comparison. In 2023, Oregon’s Senate Bill 303 directed psilocybin service center operators to report aggregated, de-identified client demographics, reasons for requesting services, and adverse reactions quarterly to the Oregon Health Authority (OHA), beginning in January 2025, (Oregon Legislature, 2023) producing the first standardized data from a supervised psilocybin service. (Oregon Health Authority, 2026) Separately, the 2024 National Survey on Drug Use and Health (NSDUH), a nationally representative survey of U.S. adults, added a psilocybin recency item, enabling the first psilocybin-xspecific past-year prevalence estimates. (U.S. Department of Health and Human Services; Substance Abuse and Mental Health Services Administration; Center for Behavioral Health Statistics and Quality, 2024)

Leveraging these two data sources (**Figure 1A**), we compared individuals accessing supervised psilocybin services in Oregon with adults reporting unsupervised use nationally across demographic, clinical, and motivational characteristics. These comparisons inform program design as additional states develop regulatory frameworks.

**Figure 1.**
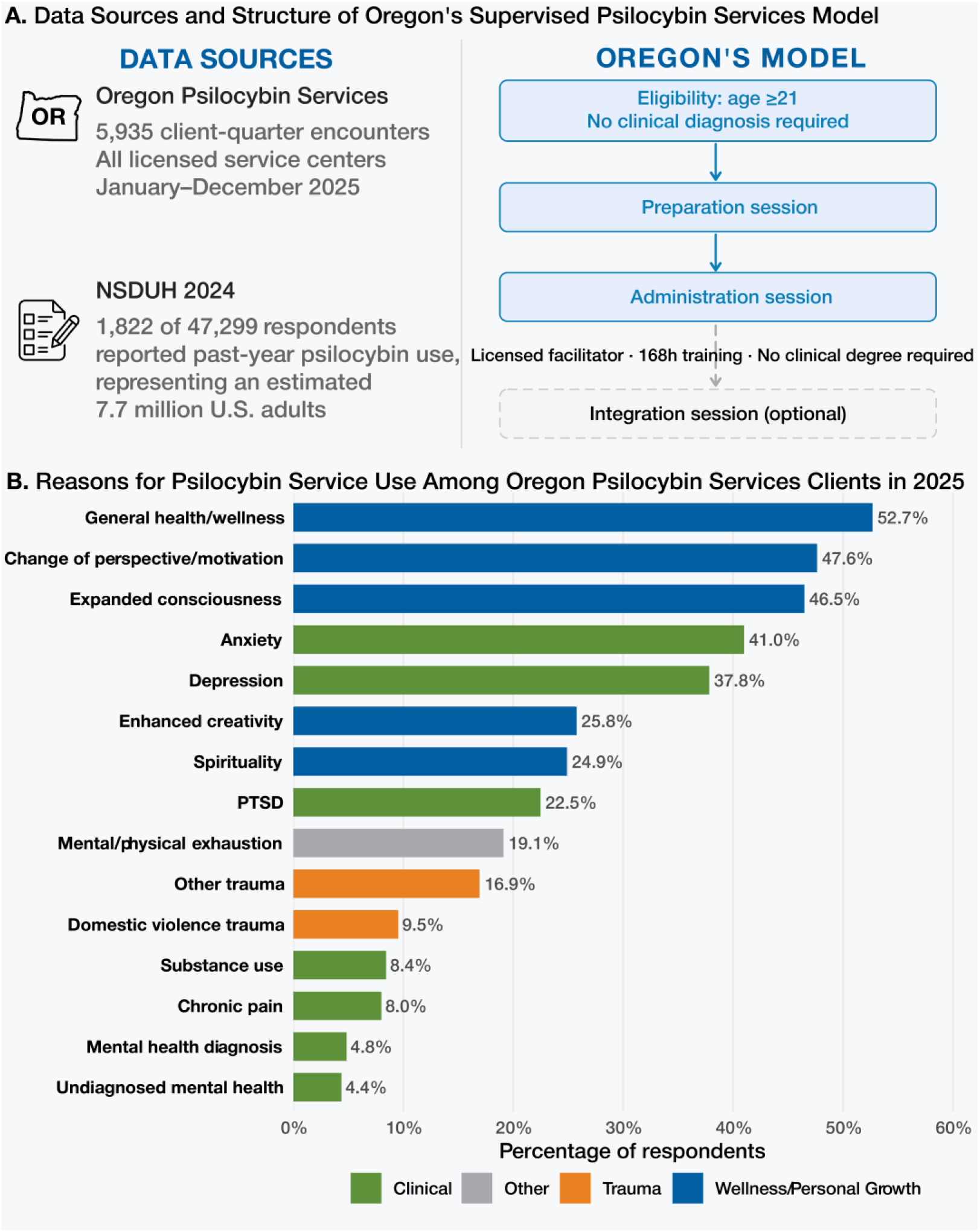
Oregon’s Supervised Psilocybin Services Model and Reasons for Service Use, January Through December 2025. Data sources and structure of Oregon’s supervised psilocybin services model. Aggregate data were obtained from all licensed Oregon service centers (5,935 client-quarter encounters, January through December 2025) and the 2024 National Survey on Drug Use and Health public-use file (1,822 of 47,299 respondents reported past-year psilocybin use, representing an estimated 7.7 million U.S. adults). Oregon’s pathway requires clients to be aged 21 years or older with no clinical diagnosis; services include preparation, administration, and optional integration sessions delivered by statelicensed facilitators who complete at least 168 hours of training and are not required to hold a clinical degree. (B) The 15 most frequently endorsed reasons for psilocybin service use among Oregon Psilocybin Services clients in 2025. Clients could select all reasons that apply; percentages are based on 3,447 demographic respondents and sum to more than 100%. Bars are colored by category: blue indicates wellness/personal growth; green, clinical indications; orange, trauma-related reasons; gray, other. Excludes “Other reasons,” “Don’t know,” and “No answer” responses.

## METHODS

We analyzed the Oregon Psilocybin Services (OPS) aggregate data from 2025 (Oregon Health Authority, 2026) (5,935 encounters across all licensed services) and the 2024 NSDUH public-use file (U.S. Department of Health and Human Services; Substance Abuse and Mental Health Services Administration; Center for Behavioral Health Statistics and Quality, 2024) (a disclosure-protected subsample of 58,633 records; 47,299 adults, of whom 1,822 reported past-year psilocybin use, representing 7.7 million adults). We applied NSDUH survey weights that account for the stratified, multistage sampling design.

Missingness ranged from 42-77% across variables (**Table S1**), due to client opt-out or service center reporting noncompliance. (Oregon Health Authority, 2026) Therefore, we calculated OPS percentages among respondents only. This respondent denominator yields higher proportions than reports calculated over all encounters. (Yu et al., 2026) All demographic characteristics were self-reported in both sources: NSDUH respondents completed computer-assisted self-interviews, and OPS clients completed intake questionnaires administered by licensed facilitators. When possible, we harmonized categories across sources (age, sex, race, and ethnicity) and compared them using two-sided chi-square tests. Income brackets differ between sources and are presented separately. OPS data also include nine self-reported disability domains consistent with American Community Survey categories.

Analyses were conducted in R version 4.5.2. Analytic code is publicly available at https://github.com/Lab-Pain/oregon-psilocybin-nsduh. This study followed the Strengthening the Reporting of Observational Studies in Epidemiology (STROBE) reporting guidelines. All data are deidentified and publicly available. This secondary analysis of deidentified, publicly available aggregate data did not constitute human-subjects research and did not require institutional review board review or participant consent.

## RESULTS

Individuals accessing supervised services in Oregon were predominantly female (58.2%) and aged 35 years or older (82.1%), whereas those reporting psilocybin use nationally were predominantly male (64.0%) and younger than 35 (56.7%). Both sex and age differed significantly between populations (p < .001). Both populations were majority White (74.3% OPS; 70.9% NSDUH). Racial and ethnic minority representation was narrower in OPS: 0.9% Black and 2.5% Hispanic, compared with 5.4% and 14.3% nationally (p < .001). OPS clients had higher incomes: 58.5% reported annual earnings exceeding $95,000 and 27.9% exceeding $182,000, whereas only 15.0% earned below $45,000. In contrast, 34.1% of adults reporting psilocybin use nationally had household income below $50,000 (**Table 1**). Most clients resided outside Oregon: 55.2% in other U.S. states and 5.8% internationally. The mean number of quarterly encounters per client was 1.28. Twenty-eight adverse events were reported (0.47%), including 9 severe reactions (**Table S2**).

**Table 1.**
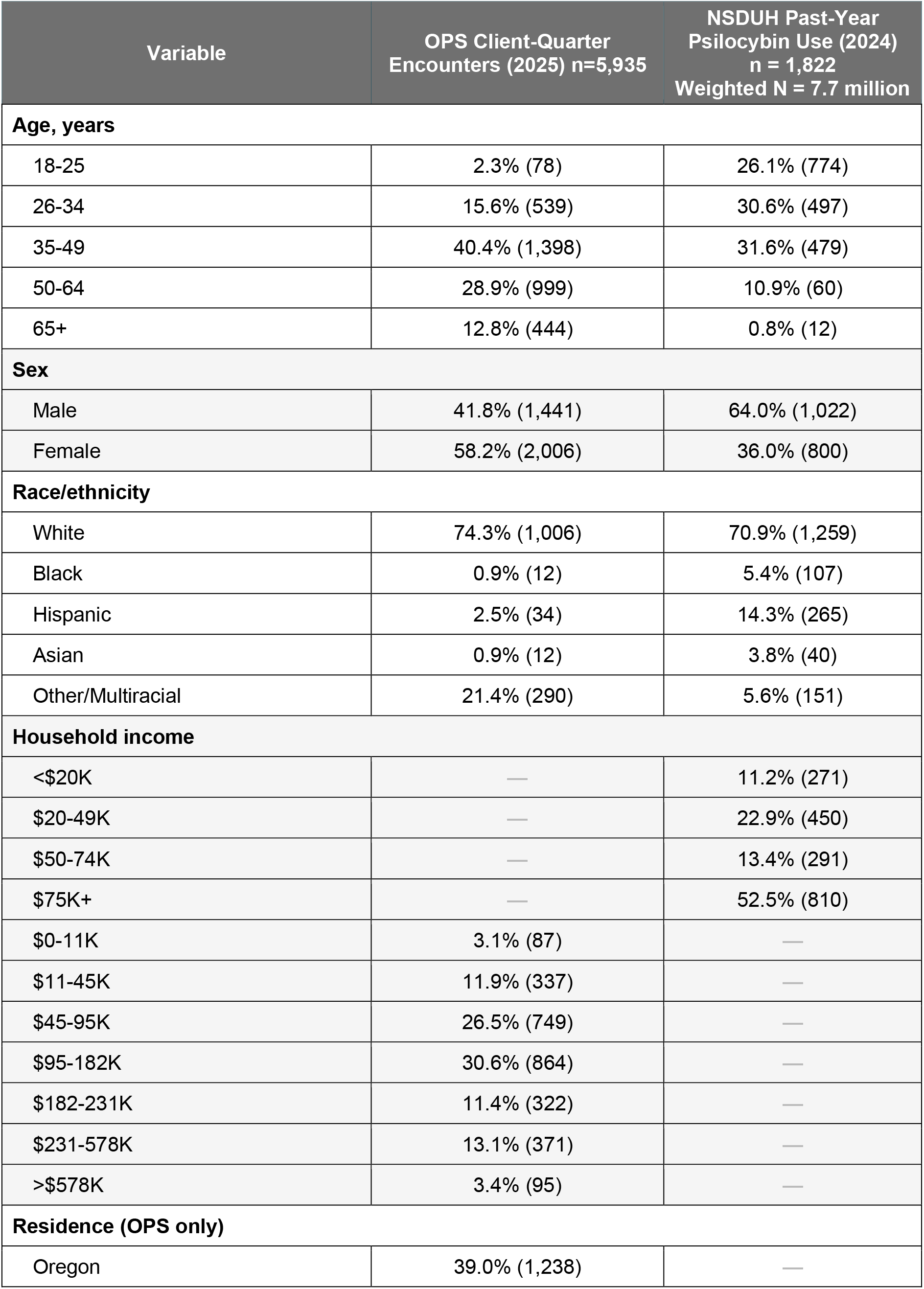
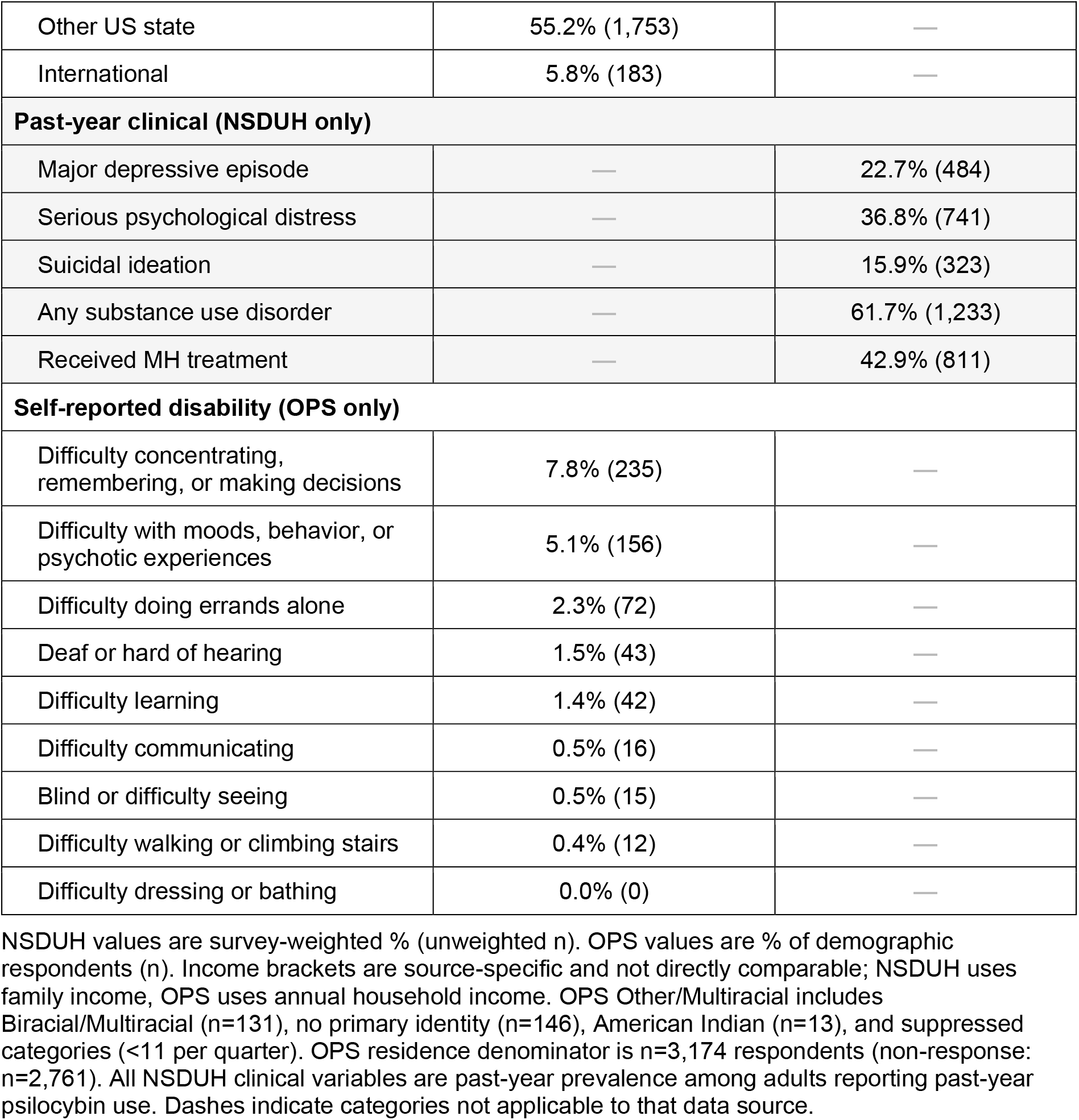
Demographic Characteristics of Oregon Psilocybin Services Client-Quarter Encounters in 2025 and Adults Reporting Past-Year Psilocybin Use Nationally in 2024.

OPS clients predominantly cited wellness-oriented reasons for accessing services: general health and wellness (52.7%), change of perspective or motivation (47.6%), and expanded consciousness (46.5%). These were followed by clinical reasons: anxiety (41.0%), depression (37.8%), and posttraumatic stress disorder (22.5%, **Figure 1B**). Self-reported functional disability among OPS participants was uncommon: serious difficulty concentrating, remembering, or making decisions (7.8%). Serious difficulty with mood, behavioral control, or psychotic experiences (5.1%), and difficulty doing errands alone (2.3%). OPS data do not include validated psychiatric measures. By contrast, adults reporting psilocybin use nationally had high rates of past-year major depressive episode (22.7%), serious psychological distress (36.8%), substance use disorder (61.7%), and receipt of mental health treatment (42.9%).

## DISCUSSION

By combining the first standardized data from Oregon’s supervised psilocybin services with a nationally representative survey of psilocybin use, we identified two distinct populations using psilocybin. Oregon’s supervised clients were older, more often female, higher-income, less racially diverse, and primarily wellness-motivated. Adults reporting psilocybin use nationally were younger, more racially diverse, lower-income, and carried substantially greater clinical burden. The low rates of functional disability reported by OPS participants contrast with the psychiatric burden observed among those using psilocybin nationally. Supervised and unsupervised psilocybin use appear to operate in parallel: one serving a narrow, clinically stable, higher-income demographic, the other a broader population with substantially greater clinical burden using psilocybin without monitoring.

Oregon’s regulatory framework helps explain why supervised services reach this narrower population. Because no diagnosis is required and services are open to any adult aged 21 years or older for any reason, (“Oregon Revised Statutes Chapter 475A: Psilocybin Services (Ballot Measure 109),” 2020) the program is, by design, as accessible to those seeking wellness experiences as to those with clinical need. Sessions are delivered by state-licensed facilitators who complete at least 168 hours of training but need not hold clinical degrees. (“Oregon Revised Statutes Chapter 475A: Psilocybin Services (Ballot Measure 109),” 2020; Oregon Secretary of State, 2026) Individual sessions typically cost $850-3,000 and can exceed $5,000, paid entirely out of pocket. Insurance does not cover psilocybin services, and subsidized access remains limited to a small number of centers and a philanthropic fund with waitlists exceeding its capacity. (Cheung et al., 2026; Luoma et al., 2025; Oregon Health Authority, 2023; Wilk, 2025) The income disparity between OPS clients and those reporting unsupervised use nationally suggests that cost filters access through this program. That most clients traveled from outside Oregon (61.0%) and averaged 1.28 encounters per client suggests episodic, tourism-like use rather than ongoing care. The low adverse event rate (0.47%) should be interpreted in this context: OPS clients were a self-selected, predominantly clinically stable population with low rates of self-reported disability, and OHA reporting definitions capture only reactions requiring emergency care or hospital transport. (Cheung et al., 2026)

Whether through supervised service centers, as in Oregon (“Oregon Revised Statutes Chapter 475A: Psilocybin Services (Ballot Measure 109),” 2020) and Colorado, or through permit-based access and decriminalization, as proposed in several other states, psilocybin availability is expanding without clinical eligibility requirements. (University of California. Berkeley Center for the Science of Psychedelics, 2026) Open eligibility does not exclude those with clinical need. Rather, the program operates outside the healthcare system. Psilocybin services are, by rule, not medical or clinical treatment: clinicians cannot refer patients into them, insurers do not reimburse them, and entry requires independent discovery and out-of-pocket payment. (Cheung et al., 2026; Oregon Secretary of State, 2026) These structural features select for wellness-motivated, higher-income clients. Policy decisions about cost subsidies, insurance integration, and clinical linkage will determine whether supervised access reaches a broader, treatment-seeking population.

These findings should be interpreted in light of several limitations. These underscore gaps in the current data collection regimes. Oregon’s mandate requires only aggregate reporting, which precludes individual-level analysis, cross-tabulation, and deduplication of clients across quarters. Missingness was substantial (**Supplementary Table S1**). Because demographic reporting is opt-out, data are plausibly missing not at random, and respondent-based estimates may carry systematic bias. Opt-out rates were nonetheless stable across quarters (33.6%-37.8%). The aggregate format precludes imputation, which requires individual-level records. The public-use NSDUH file contains no state identifiers, precluding an Oregon-specific comparison, (Substance Abuse and Mental Health Services Administration, 2025) because 61.0% of encounters involved clients residing outside Oregon, a national comparator remains the more appropriate frame. States developing new psilocybin frameworks should mandate individual-level, linkable data collection and incorporate brief standardized outcome measures to evaluate whether services are beneficial. The NSDUH, by contrast, does not capture the source, setting, or supervision context of psilocybin use, precluding assessment of whether legalization diverts use from unregulated to regulated channels.

The demographic profile of individuals accessing Oregon’s supervised psilocybin services differs substantially from that of adults reporting psilocybin use in the general U.S. population. Oregon’s open-access model predominantly serves higher-income adults seeking wellness while millions with substantial psychiatric comorbidity use psilocybin unsupervised. Thus, the safety and utilization data generated by the current system reflect this self-selected sample and should not be generalized to the broader population seeking psilocybin therapeutically.

As more than 30 states consider similar open-access frameworks, (Siegel et al., 2023; University of California. Berkeley Center for the Science of Psychedelics, 2026) cost structure, insurance coverage, and clinical referral pathways will determine whether supervised access expands beyond a wellness-seeking demographic. Broadening who supervised services reach and generating outcome data relevant to populations currently using psilocybin without clinical monitoring should be central design priorities.

## Data Availability

All data analyzed in this study are publicly available and deidentified. The 2024 National Survey on Drug Use and Health public-use file is available from the Substance Abuse and Mental Health Services Administration (https://www.samhsa.gov/data/data-we-collect/nsduh-national-survey-drug-use-and-health/datafiles/2024). Oregon Psilocybin Services quarterly aggregate data files are available from the Oregon Health Authority data dashboard (https://www.oregon.gov/oha/ph/preventionwellness/pages/psilocybin-data-dashboard.aspx). Analytic code is publicly available at https://github.com/Lab-Pain/oregon-psilocybin-nsduh.

https://www.samhsa.gov/data/data-we-collect/nsduh-national-survey-drug-use-and-health/datafiles/2024

https://www.oregon.gov/oha/ph/preventionwellness/pages/psilocybin-data-dashboard.aspx

## Acknowledgments

None.

## Study Type

Cross-sectional study.

## Data Sharing Statement

The NSDUH 2024 public-use file is available from the Substance Abuse and Mental Health Services Administration (https://www.samhsa.gov/data/data-we-collect/nsduh-national-survey-drug-use-and-health/datafiles/2024). Oregon Psilocybin Services quarterly data files are available from the Oregon Health Authority data dashboard (https://www.oregon.gov/oha/ph/preventionwellness/pages/psilocybin-datadashboard.aspx). Analytic code is publicly available at https://github.com/Lab-Pain/oregon-psilocybin-nsduh.

## Notes

### Competing Interest Statement

Financial competing interests (past 36 months): JPD has received medication provisions from Jazz Pharmaceuticals and Ananda Scientific and has been a compensated consultant for Boehringer Ingelheim. CP has consulted for Freedom Biotechnology, Mind Therapeutics, and Biohaven Pharmaceuticals; has received research funding from the Heffter Foundation, the Steven and Alexandra Cohen Foundation, the Taylor Family Foundation, Transcend Pharmaceuticals, Ceruvia Life Sciences, Freedom Biotechnology, Biohaven Pharmaceuticals, the National Institutes of Health, the Brain and Behavior Research Foundation, the Colton Center for Autoimmunity at Yale, and the Department of Defense / Congressionally Mandated Research Programs; is an inventor on patents related to novel psychedelic drug combinations, autoantibodies implicated in neuroimmune pathogenesis, and novel pharmacotherapies for OCD, Tourette syndrome, and related conditions; and has received royalties from Oxford University Press, UpToDate, Biohaven Pharmaceuticals, and Ceruvia Lifesciences. GPAC and CR report no financial competing interests. Employment, stocks, and other paid activities (past 36 months): CP holds equity in Mind Therapeutics, Alco Therapeutics, Loop Bio (pending), and Lucid/care (pending). No author has been paid to serve as an expert witness on the subject of this study within the past 36 months. The other authors report none. Non-financial competing interests: None.

### Author Declarations

The study used ONLY openly available, deidentified human data that were publicly released before the initiation of the study: (1) Oregon Psilocybin Services quarterly aggregate data files (2025 Q1-Q4) published by the Oregon Health Authority on its public data dashboard, https://www.oregon.gov/oha/ph/preventionwellness/pages/psilocybin-data-dashboard.aspx; and (2) the 2024 National Survey on Drug Use and Health public-use file released by SAMHSA, https://www.samhsa.gov/data/data-we-collect/nsduh-national-survey-drug-use-and-health/datafiles/2024. No application, registration, or data-use agreement was required to access either source.

### Funding

- Connecticut Department of Mental Health and Addiction Services

