## Supplementary Material (Tables S1-S2) for "Who Is Reached by Supervised Psilocybin Services? Oregon Services Versus National Psilocybin Use"

**Supplementary Material for *Who Is Reached by Supervised Psilocybin Services? Oregon Services Versus National Psilocybin Use***

**Table S1. Missingness of Demographic Variables in Oregon Psilocybin Services, 2025**

| Variable | Respondents, n | Missing, n | Missing, % |
|---|---|---|---|
| **Age** | 3,458 | 2,477 | 41.7 |
| **Sex** | 3,447 | 2,488 | 41.9 |
| **Residence** | 3,174 | 2,761 | 46.5 |
| **Household income** | 2,825 | 3,110 | 52.4 |
| **Race and ethnicity** | 1,354 | 4,581 | 77.2 |
| **Visit reasons*** | 3,447 | 2,488 | 41.9 |
| **Functional disability (9 domains)†** | 2,867–3,200 | 2,735–3,068 | 46.1–51.7 |

Total encounters, N = 5,935. Missingness reflects a combination of client opt-out and service-center reporting noncompliance. Denominators are variable-specific; Oregon's Senate Bill 303 mandates aggregate, non-linkable reporting, and does not permit across-variable linkage or across-quarter deduplication of clients. *Visit reasons are reported as select-all-that-apply; the denominator is approximated using sex-reported respondents as a proxy for encounters with any demographic data recorded. †Functional disability is reported separately for each of nine domains consistent with American Community Survey categories; respondent denominators vary by domain (range shown).

### Table S2. Service and Safety Characteristics of Oregon Psilocybin Services in 2025

| Characteristic | Value |
|---|---|
| **Service volume** | |
| Client-quarter encounters | 5935 |
| Individual administration sessions | 86.1% (4,628) |
| Group administration sessions | 13.9% (747) |
| Total sessions | 5375 |
| Mean psilocybin dose, mg | 24.5 |
| Mean encounters per client | 1.28 |
| **Adverse events, No. (% of encounters)** | |
| Adverse behavioral reactions | 0.10% (6) |
| Severe adverse behavioral reactions | 0.12% (7) |
| Adverse medical reactions | 0.22% (13) |
| Severe adverse medical reactions | 0.03% (2) |
| Post-session reactions | 0.17% (10) |
| Total adverse events | 0.47% (28) |
| **Service denials, No. (% of denials)** | |
| Inconsistent with business model | 36.4% (94) |
| Client ineligible | 57.0% (147) |
| Client intoxicated | 0 |
| Concerning behaviors | 3.1% (8) |
| Other reasons | 10.1% (26) |
| Total denials | 4.3% of encounters (258) |

Data from Oregon Health Authority, Oregon Psilocybin Services data dashboard, January through December 2025. Mean psilocybin dose is the average of quarterly service-center-computed means. Adverse events include reactions reported during or after administration sessions. Service denials are reported by reason; categories are defined in Oregon Administrative Rules 333-333.
